# Availability of Essential Medicines During the COVID-19 Pandemic: A Qualitative Study Examining Experiences and Level of Preparedness in Kenya

**DOI:** 10.1101/2023.10.10.23296845

**Authors:** Joseph Odhiambo Onyango, Dosila Ogira, Gilbert Kokwaro

## Abstract

This study investigated the impact of the COVID-19 pandemic on essential medicine availability in Kenya’s health system. Key informant interviews were conducted, and the data were analyzed using NVIVO software. Six themes emerged, aligning with the WHO health system building blocks. These themes provided insights into the experiences, challenges, and opportunities regarding essential medicine availability during the pandemic. The initial response involved reallocating resources, affecting the procurement of essential medicines at national and county levels. To enhance preparedness, investments are crucial in strengthening financial systems and policies, improving supply chain resilience, and promoting local production through regulatory enhancements. These strategies aim to build resilient health systems and self-reliance, particularly for countries transitioning from donor aid. The findings underscore the importance of effective preparedness to ensure the availability of essential medicines during emergencies like the COVID-19 pandemic

## Introduction

On January 23, 2020, the World Health Organization’s International Health Regulations (IHR) Emergency Committee, in response to the new coronavirus first detected in Wuhan China, convened and issued a directive that: “all countries should be prepared for containment, including active surveillance, early detection, isolation and case management, contact tracing and prevention of onward spread of 2019-nCoV infection, and to share full data with WHO”(1). Based on recommendations from the Committee, the virus was declared a public health emergency of international concern on 30th January 2020 (2).

One of the areas that was primarily affected by the pandemic is the availability of essential medicines. Previous studies on epidemics and pandemics have reported an increased rate of infections due to disruption of medical supplies and products such as during the 2014 West Africa Ebola outbreak (3). Similarly, the coronavirus was initially seen to negatively affect supplies in both high- and low-income countries. For instance, scholarly investigations undertaken to ascertain the impact of COVID-19 in high resource settings such as the United States and Saudi Arabia revealed shortages of ten or more essential medicines at the onset of the pandemic (4)(5)(6)(7). Chloroquine and hydroxychloroquine were some of the medicines enlisted since they were believed to alleviate the COVID-19 symptoms and reduce its severity (8). In most instances, these shortages were majorly attributed to the disruption in the global pharmaceutical supply chain (9).

Most low and middle income countries experienced a greater risk of medical product shortages due to their already weak health systems and low resource setting than HICs (10). These shortages were further exacerbated by low production capacity and heavy reliance on developed countries for raw materials and finished products (11). For instance, countries like China and India who have long manufactured and exported active pharmaceutical ingredients to LMICs had reduced their production capacities (12). Additionally, mitigation measures to control infections globally resulted in lockdowns and limited movement of people and products, affecting the supply of essential medicines (13).

As it stands, the Kenyan health system is still grappling with opposing needs both from communicable and noncommunicable diseases (14). Coupled with this is the Country’s high dependency on donors to fund some of its health products or interventions, amidst the donor transition, further strains the system (15). The COVID-19 pandemic saw most countries, including HICs, focussing on their needs hence LMICs were forced to look for alternative ways of meeting their needs. To depict this, early findings in Kenya indicated shortages in essential medicine including the Internationally Controlled Essential Medicines (ICEMs)(16). However, to maintain the access and supply of essential medicines to patients during the pandemic, an innovative supply-chain strategy was developed in rural western Kenya. This was done through a revolving fund pharmacy model that capitalised on stocking essential medicines to constituent health facilities that was easily accessible to patients (17).

Universal Health Coverage (UHC) is one of the main pillars of the ‘Big Four’ agenda in Kenya, however, the bid to achieve it was strained by the COVID-19 pandemic (18). For instance, KES 11.3 billion was reallocated from the UHC reservoir to boost the COVID-19 kitty at the beginning of the pandemic risking other aspects such as availing essential drugs (19)(20). Therefore, in order to ensure the availability of essential medicines and achieve UHC amidst other competing healthcare needs, there is need for innovative solutions to match especially during unprecedented times (21). This study sought to understands how the COVID-19 pandemic has impacted the access to essential drugs in Kenya and some of the actions stakeholders can take to improve country preparedness for future pandemics. Maintaining continued health progress will depend on how the health policymakers in the world navigate this pandemic through their emergency preparedness.

### Study objectives

The main aim of this study was to understand how the COVID-19 pandemic has affected the availability of essential medicines in Kenya. Specifically, we looked at the health systems gaps that challenged the access and provision of essential medicines. Additionally, we explored opportunities that can be leveraged to improve the country’s preparedness for future pandemics.

## Methodology

### Study design and sampling technique

This study employed a cross-sectional qualitative design to meet its objectives (22). Additionally, due to its qualitative nature, the participants were selected purposively, focusing on those who had experience in the sector and were willing to share the information. Interviewees were asked about their knowledge and experience with the availability of essential medicines during COVID-19. This study was conducted during the surge of COVID-19 from July 2021 and was completed in July 2022.

### Data acquisition

We integrated desk review and key informant interviews to collect data and achieve the study objectives. For the desk review, a comprehensive review of grey and peer-reviewed literature on the impact of COVID-19 on essential medicines was carried out sequentially from the global, regional, and local contexts to better understand and explain the phenomenon. For the key informant interviews, we carried out 20 semi-structured interviews. These included representatives from; (i) National government-2, (ii) Sub-national County government-7, (iii) Health professional bodies-2, (iv) Pharmaceutical manufacturers-5, (v) NGOs and CSO representatives-4. Due to the initial movement restrictions resulting from the COVID-19 pandemic, the interviews were carried out virtually after obtaining oral consent from the participants.

### Data analysis

We used framework approach for analysis (23). The analysis team began by reading through all the transcripts which had been transcribed verbatim. After the data familiarization, an initial codebook was developed drawing on the study objectives and integrating emerging themes from the interviews. After refining the codebook, the interviews were coded using NVIVO software. The findings were analysed and presented based on the prevalent views.

### Ethical consideration

This study sought all the ethical approvals and research permits. The study was conducted virtually due to the social distancing measures from COVID-19, and all participants were requested to consent verbally. A study information sheet contains information on; 1) the title of the Study and why it is being carried out, 2) which participants were going to be involved and why, 3) time demands for the participants, 4) the voluntary nature of participation into the Study, 5) assurance of confidentiality once they agree to participate and 6) how the data provided will be utilized; was shared with participants on email a few days before the interview was set. All recordings and transcripts were deidentified to protect the interviewee’s confidentiality.

### Findings

The findings are categorized into six broad themes outlining the experiences, challenges, and opportunities in relation to the availability of essential medicines during the COVID-19 pandemic in Kenya. These include (i) healthcare financing of essential medicines, (ii) supply chain and procurement system for essential medicines, (iii) regulatory policies and manufacturing capacity of essential medicines, (iv) human resource for health training on production of essential medicines, (v) health information system on essential medicines, and (vi) the role of the public versus private sector in accessing essential medicines and promoting UHC.

### Healthcare financing for essential medicines

Most of the respondents reported that the initial reaction by the Country involved the revision of priority gaps which resulted in the diversion of resources, including financial resources towards management of the COVID-19 pandemic. For instance, it was mentioned that the price of some non-pharmaceutical commodities used for containment, such as surgical gloves and masks went up almost 20 times, and more funds had to be allocated towards their purchase. This priority shift was seen to affect other aspects in the healthcare sector including the procurement of essential medicines both at the National and County levels.

> “So, there was really a big shift in terms of the Country’s focus, and I’m not only speaking about Kenya, I am also speaking about Africa in general. We saw the regulators and the health authorities, shifting their focus and even stopping and forgetting about the essential medicines and now trying to focus on COVID. Initially, there was lack of balancing out, to ensure that the essential medicines are still being accessed, while they are still managing a pandemic.”

> Key Informant 2

> “So, pre-corona we could supplement well what KEMSA didn’t have. But Corona came in and things went upside down. All the County funds that were meant for essential commodities were devised to buy preventive commodities for coronavirus and it meant that now we had less commodities that were always available with us. So, it meant fill rate prescriptions and essential pharmaceutical availability went down.”

> Key Informant 7

Some respondents faulted the Country’s emergency preparation due to lack of financial reserves dedicated towards emergency response. This was seen to slow down organic and coordinated response including acquisition of essential medicines. However, as the pandemic progressed, some counties called for emergency meetings to reallocate funds for purchasing of some of the essential medicine that were running out of stock then. Additionally, high level of donor dependency for some essential medicines was also outlined as a challenge and if there was a reduction, this was seen to affect the flow of essential medicines.

> “…we have an ineffective response due to the inadequate and lack of a dedicated county resource kitty towards emergency response. So of course, when I talk about emergency response, you will also have essential medicine as an aspect of the emergency response. There is nothing that exists currently.”

> Key Informant 6

> “We rarely set funds for emergency waiting. And you find most of the time we don’t have those contingency – our contingency and emergency preparedness is not strong. We tend to work at the spot, and I think that’s also what we did in the COVID emergency”

> Key Informant 12

> “… for the big diseases, HIV, TB, malaria, it’s still largely donor-dependent, which is not sustainable. And I mean, that’s even what led to the shortage of ARVs because rather than making them locally or procuring for ourselves, we were waiting for donor supplies – if you are completely donor-dependent, then you can run out of products if the donor doesn’t send you money, if donor says your procurement systems are corrupt or they are not up to standards they’ll not send products.”

> Key Informant 19

Additional recommendations to be adopted to alleviate challenges facing financing of essential medicines in the event of a future pandemic were made by respondents. First, some respondents expressed a need for improving financial policies through establishing facility improvement funds and financial reservoirs that could be retained at the facility level. This was seen as a sustainable way of ensuring that commodities are available and can act as a buffer in the event of a pandemic and increase the accessibility of medicines to patients. Secondly, suggestions were made to improve financial forecast to match up with the financial allocation of essential medicines especially in the public sector in the Country. This was seen as ensuring access to essential medicines hence promoting Universal Healthcare Coverage (UHC).

> “So, we should start building up some kind of funding for any eventuality or calamity that might be used for essential medicines – we don’t have that kind of funds sitting for free for such kind of scenarios”

> Key Informant 11

> “One of the recommendations is to ensure that the public sector increase allocations for essential medicines and that they are monitoring the needs versus the expenditures for the medicines. That is a critical one especially for sustainability of UHC…following protocols, following the essential list and ensuring that allocations are based on established needs.”

> Key Informant 15

### Supply chain and procurement systems for essential medicines

During the COVID-19 pandemic the supply chain system was heavily disrupted globally. Respondents mentioned that the rigidity of movement attributed by measures such as lockdown slowed down the production and access to the essential medicines. For instance, manufacturing countries like India and China that produce active pharmaceutical ingredients and supply Kenya had highly reduced their exportation rate, with some factories shutting down because of COVID-19 cases, negatively impacting production of essential medicines. Additionally, with reduction in production, there was also an increase in competition for raw materials and a subsequent increase in cost of essential medicines across countries due to high demand and low supply.

> “Yes, essential medicines were impacted. And this was mainly because of the manufacturing dependency we have from the global aspects, whereby most of our companies manufacture in a different facility and therefore, at each center of the globe, there was an impact in terms of manufacturing. There was a shortage of the Active Pharmaceutical Ingredient, a lot of logistics problems, and of course, there was restricted movement, so therefore, restricted movement of essential medicines.”

> Key Informant 1

> “The answer is yes, there was a difference in cost of acquisition pre and post pandemic. There was generally high cost of medications because of transportation cost going up. This was affecting general medicine because some of our courier companies that we use for transportation of medicines closed shops. With that cost was going up so a basic medicine that you would need maybe to get with 100 Kenya shillings (Approximately 1USD), we were forced to part with around 150 Kenya shillings simply because of the transportation cost.

> Key Informant 17

Almost all respondents from sub-national level reported that there was deviation from what was a routinely smooth supply chain and procurement system cascading from the national government with extended lead time. With many efforts being redirected towards the containment of the pandemic, it was mentioned that shortages were experienced even for some of the key essential medicines such as paracetamol. However, some respondents noted that not all essential medicines were affected the same way and some of the shortages were artificial due to consumers overreliance on brand prescription and the preceding prescribing pattern.

> “When COVID came, it ended up interfering with our cycles of replenishing stocks. In this stock-out, there was artificial shortage in particular drugs as simple as paracetamol tablets. We’ve had very serious shortages across the year 2020 and part of 2021.”

> Key Informant 5

> “Some patients only believe in the products that’s written on their prescription and they were not willing to switch maybe to another brand or to a generic …that’s what I mean by artificial shortage, some patients on blood pressure medication would feel that the products they’ve been on for so long is not there and it’s difficult to switch them to a similar product, but another brand or a generic.”

> Key Informant 15

However, as the pandemic progressed, the government took swift measures to alleviate the hurdles faced by the supply chain. For instance, in terms of limited transportation of essential medicines due to lockdown, the government issued permits to distributors who were considered essential workers. Additionally, other counties leveraged previous good working relationships with local private suppliers, which helped them acquire critical essential medicines at the time. Other innovative ways of strengthening the supply chain were also witnessed, for instance, some counties restructure their systems to distribute essential medicines to smaller facilities. This was seen to help patients access essential drugs within their locality without travelling longer distances.

> “We had to work with our suppliers, there are those who we are having a good rapport with them and despite the business environment at the time, they were able to supply us – we were able to negotiate, and they supplied us when we did not have funds as they waited for us to process those funds.”

> Key Informant 7

> “Then also we are coming up with a pilot which we are still in our initial stages of implementation, this patient who used to travel to the bigger health facilities to attend their clinics and get their drugs refilled, we are now doing the other way round; we are sending drugs to these smaller facilities and also accompanied with health workers who have experience with dealing with the kind of diseases or rather the noncommunicable diseases so that this patient doesn’t have to travel again to long distance to go to the bigger facilities.”

> Key Informant 19

Recommendations were also made to ensure that there is always focus on the supply and access of essential medicines. For instance, it was mentioned that the supply chain systems should be robust by setting up real-time systems, providing a good quality management system process and encourage stockpiling to ensure that supply and access of essential medicines is maintained.

> “We’ve also noticed that we need to have huge buffers for some of our medicines. We need to have buffer stocks and if possible, have county stores that can really help us in terms of emergencies how to move commodities.”

> Key Informant 5

### Regulatory policies and manufacturing capacity of essential medicines

Almost all the respondents faulted the government for having rigid regulatory policies and systems not responsive to unique situations such as the pandemic. Despite being an emergency, it was observed that the Country took time to introduce alternatives for outsourcing essential medicines or giving approvals, which would have improved the response rate and interventions at the time. Additionally, there were initial cases of corruption witnessed at the Country’s main medical supplier to counties, which negatively affected the stock levels and flow of essential medicines.

> “Yes, I would say due to our procurement system at the County level, including that of essential medicines, we are restricted to where we can procure. We procure from KEMSA. Going out of that system we are breaking the law. So maybe a policy direction can be added to guide us if these entities do not have any commodities such as during COVID, then legally you can procure from any other source.”

> Key Informant 18

> “The fact that there had been no emergency use authorization guideline and there was really no way of managing a situation like this, resulted in a lot of time wastage. Because the regulation had not been implemented, had not been developed, nor had it been structured…if I speak for Kenya, you find there is a lot of back and forth, whereby some quick acceptance of the stringent regulatory authorities and mutual recognition would make that process much easier.”

> Key Informant 12

> “And also, you know, for us we work within the county so our main supplier is the Kenya Medical Supplies Agency, KEMSA, and with whatever happened, the gaps in accountability, it messed up the stock levels and as a result, those of us who depend on that particular body as the main supplier were directly affected. And as I said overall there was that decrease in supply.”

> Key Informant 6

It was also noted that the government policies have not been coherent or conclusive of promoting local manufacturing of essential medicines despite the Country’s ability and capacity. Some respondents pointed out that there were gaps, including unfavorable tax brackets for the local manufacturers leading to high manufacturing costs. As a result, respondents felt that this created unfair competition for the local manufacturers. Additionally, the Country was seen to heavily rely on importation of active pharmaceutical ingredients to manufacture finished products, contributing to the shortage of essential medicines during the pandemic.

> “We talk about India, we talk about Bangladesh, why do we keep on talking about these two countries? Why can’t we not do it here?… And we have more than 30 manufacturers, more than any country between the Maghreb countries and South Africa, more. And we’re losing an opportunity for Kenya simply because, policy makers, decision makers are not aligned, they’re going to miss the golden opportunity that is there.”

> Key Informant 13

> “But let’s go back to the essential medicines, there are several essential medicines I don’t manufacture. And the reason is, that it doesn’t make it viable when essential medicines are being imported from India, Pakistan, China.”

> Key Informant 18

> “The government decided that we go from zero rated to exempt, but they don’t understand the fundamental difference between the two in the VAT context. When we were zero rated, whatever services we paid for electricity, consultancy etc., we could claim the VAT back. However, when it’s an exempt, we’re unable to claim this. So, what it means is that about 3% to 4% now come as a cost to our products. But when you import things from UK from India, it does not affect them. So, you’re automatically making an unleveled playing field for your local manufacturers.”

> Key Informant 16

Amidst the challenges, some respondents noted that there was progressive response from the government which saw introduction of regulatory approaches aimed at strengthening and supporting local production of essential medicines.

> “Right now, if you go to the national treasury there is a finance bill speaking to aspects of exemptions and tax reliefs to local manufacturers to be able to start manufacturing in the county.”

> Key Informant 12

Some respondents felt that improving local manufacturing capacity for the Country to be more self-reliant would have sealed the gap and improved the shortage of essential medicines. Expanding the manufacturing portfolio even to include active pharmaceutical ingredients that produce various finished products was seen as one of the ways.

> “Self-reliance and sustainability are critical for a country. Before, we would speak of one global village where you can produce in China, assembly in India, use in Europe or Kenya. COVID-19 has challenged that in terms of you need some slight level of self-reliance. If you are not producing anything, you are at a risk.”

> Key Informant 9

> “If you look at the essential list, you’ll find a lot of the big manufacturing sites here in Kenya are manufacturing only three or four elements, mainly the analgesic portfolio, where you have your paracetamol, and a few of the anti-infective’s where you have your antibiotics being produced. So, there is need to scale up local production and local manufacturing to a higher level whereby we reduce that reliance to external manufacturing.”

> Key Informant 2

Other recommendations included strengthening the regulatory authority to encourage foreign investors to venture in production in the Country and incentivising local manufacturers through improving tax policies to reduce cost of production. Strengthening multi sectorial and cross-country ties was also highlighted to improve the availability of essential medicines.

> “We have to spur – we have to give financial incentives, tariff and non-tariff incentives for local manufacturers to manufacture more. One, to manufacture at a higher capacity, number two, for expansion into more complex molecules on the essential medicines list like vaccines and other products.”

> Key Informant 17

> “So, in terms of even formulating strategies or putting bilateral agreements, governments to governments, Country to Country is a very important factor that we should be looking at. Do we have the capacity to manufacture raw materials? No… So how we can better engage and build our relationship with the countries that are able to afford and offer us those raw materials is what we should be looking at as a government in terms of policy decisions.”

> Key Informant 8

### Human resource for health training on the production of essential medicines

> Few of the respondents pointed out a gap in the curriculum and training in the Country, especially around the production of essential medicines. They felt that the current curriculum had limited content in the production of complicated molecules, and there was need to expand it and spur more manufacturing even into human vaccines, some of which are on the essential medicines list.

> “We are not manufacturing any human vaccine, so that policy issue around more complex manufacturing even ties in with training institutions and the training program for chemists, pharmacists, and biomedical engineers. We definitely need to have a skill lab so that people have a mindset to set up plants of their own and they know what to do.”

> Key Informant 16

> “The other area is the area of training, and this is another area that really is missing especially looking at the impact of the regulatory science in this and industrial pharmacy. The career of the pharmacists needs really to be tailored so that it is in line to with these requirements.”

> Key Informant 4

However, one of the respondents mentioned that the government had initiated strategies to incorporate these aspects of training. They illustrate;

> “And for XXX, yes, we are starting. And in fact, we’ve started as an in-house, we’ve developed courses for training of our recruits that come in…we’ve developed it as an online system so eventually, we are hopeful that we shall be able to stimulate the expansion of the regulatory science into universities because it will also really help.”

> Key Informant 14

Recommendations were made on building the capacity of healthcare professionals through targeted training programs on production of essential medicine. This was seen as a sustainable way of addressing the needs of the Country in the event of a future pandemic. Some of the respondents illustrate;

> “We need to look at the developments happening in the world and what we can do as a region. Build on our capacities, build on our manufacturing knowledge through training of our HRH…build on it so we become a net export of pharmaceuticals to Africa.”

> Key Informant 3

> “So, the other thing is increasing the number of human resource because that is also another area. The regulator is required to strengthen research in the Country, and it cannot strengthen research in health products and technologies if it has limited staff. So, we need to strengthen the research within the Country and when you’ve got that strengthening of research, we will be able to link the industries with the universities and research institutes for development of essential medicines.”

> Key Informant 18

### Health information system on essential medicines

It was noted that existing information system was not robust and there were instances of downtime which affected the ordering of essential medicines. Additionally, process documentation for future forecasting and quality management system was also seen as a point of weakness both at the national and County levels.

> “In terms of the procurement and information system, as I mentioned, there had been a slowdown, based on the lack of the systems being independently run. There is a lot of dependence on individuals rather than the system, so this means if that individual is unwell then the process stalls.”

> Key Informant 13

> “I also think a big gap that we have there is documentation. We are not really strong in terms of documentation real-time of what is happening, and interventions being made. And then you know right now when things are ‘normal’ we are meant to have done like a post epidemic sort of postmortem to see whether the interventions worked, or they didn’t work and where we could improve to ensure access of essential medicines. I mean, that’s how we make our whole system better – supply chain and even the larger response system. But we don’t do that as a country neither do we do it as a county.”

> Key Informant 6

However, few respondents observed that the pandemic created opportunities for improvement of the health information system concerning availability of essential medicines. For instance, the pandemic heightened the need to improve and strengthen supply chains through digitization. Additionally, in some instances there was a move from manual to automated database systems that can be used to forecast the use and need of essential medicines in the Country.

> “There was a lot of improvement towards supply chain and even digitalization… you heard of a system called ‘**chanjo’** (immunization), it was actually conceived to manage vaccines and the supply chain.”

> Key Informant 12

> “…you can imagine managing something like paracetamol, so many tablets in a day, you can dispense so many. In a manual system it’ll be so hard for you to even manage like those consumption data but right now there is a lot of commitment by government to put up digital system led to early visibility for supply chain.”

> Key Informant 14

> “The other area is for us is our databases; you may be aware that we moved all our activities to online activities and our system is working but we are improving on it. And what we are going to do, we’ve got a repository of all the registered and retained products, including essential medicines, into a system and from that system which we are still evaluating applications that have been done into it so that we can review both current and historical data. The next thing of course is to make this data dynamic and one of the aspirations is to have a system that would have really track and trace capabilities.”

> Key Informant 4

Recommendations were made to set up solid, secure systems with a clear quality management system protocol, ensuring that despite situations of calamities or pandemics, the systems continue to run. Additionally, respondents reported a need for better data collection and documentation improving data collection that can be used for fact-based decision making.

> “And also, the fact that, when there is the move into a digital space, that also would have really helped the process. Because where you would be expected to go to the Ministry of Health to follow up on something and of course you find there is a COVID restriction, there is no movement, and the officers are not there, nothing happens. So, moving, adopting more of the digital platforms, which I think slowly over time they have been able to do that.”

> Key Informant 15

> “As a country, we need to improve on our surveillance measures. It will help us discover new things and maybe now use this data to prevent other pandemics that might come up in future and the need for more evidence-based decision making from whatever we collect and even project the medicines we need.”

> Key Informant 10

### The role of the public versus private sector in accessing essential medicines and promoting UHC

The respondents highlighted the long-standing gaps in availability of essential medicines in the Country’s public health system which were alleviated by the COVID-19 pandemic. Some of them attributed this to the inefficiencies in supply chain forecasting in the public sector which in some instances has resulted in expiries and stock outs of essential medicines. They believed that harmonization of the public and private supply chains could reduce the mismatch and improve availability of essential medicines.

> “But most importantly is that there is a general lack of supplies within our public system but that now makes it necessary for patients to access the same medicines in the private sector and at higher prices. So, during COVID the same situation was either worsening for some medicines depending on their demand.”

> Key Informant 12

> “In terms of downstream manufacturing, I would say in the private sector that is sorted. You know that usually flows because its market driven and so it’s quite efficient. But in the public sector we don’t see that type of efficiency and that’s why we see expiries, stock-outs and poor storage at KEMSA. So, for downstream public sector, we just have to strengthen KEMSA because now counties must buy from them by law. So, we just have to strengthen their forecasting and their good distribution systems.

> Key Informant 14

Some of the respondents, majority of who were from the public sector, highlighted the opportunities for collaboration with the private sector to ensure continuity of flow of essential medicines. They noted that the private sector tried to bridge some of the gaps that the government was unable to sort out during the COVID-19 pandemic.

> “Okay, I think that the private sector has tried to support us. Some gave out masks and general PPEs for our CHVs and the community at large and this they were providing, and they were intending to do this for a whole 12 months starting somewhere in January. Then with regards to our private chemists, they’ve been the middleman who ensures that commodities are available. Though its business but at least they’ve made sure that whenever government has gaps in terms of commodities at least the private sector has drugs that they can get.”

> Key Informant 5

> “Yes, to some extent we still have some of our stakeholders being private entities and mainly those who supply us with medicines through the tender system. They came in handy because when we were low on finances, we were able to negotiate with them and they supplied us on a debt.”

> Key Informant 7

> “Some of the partners, if we knew they were going to a particular county, we would tell them to take these commodities deliver them to the Sub County hospital from where now the facilities would pick. So, we rode on their mobility because they had vehicles, they had fuel and are always moving across. So, I would say in terms of that collaborating aspect between the NGOs and the private sector, the main area they really helped us in was supplies.”

> Key Informant 18

There was recognition and consensus among respondents that the private sector played a very important role in terms of promoting availability of essential medicines. Recommendations were brought forth to foster collaboration between the public and private sector as it was seen as the most sustainable way of achieving UHC even during unprecedented times.

> “And it’s very evident that public cannot go alone at it, if you’re talking about the supplier, the big suppliers are coming from the private manufacturing companies. So, without that collaboration there is no way that the government will eventually gain from these if they move at it alone.”

> Key Informant 2

> “Okay, so there is my number two takeout, is collaborating across the centers, whereby we really need to strengthen the private-public partnership, and this means diversifying not only with the government, not only whereby we are working together, non-government organizations…the civil society, having everybody in and hearing everybody’s voice, whereby we end up having a unified approach towards this. So, collaborating across the sectors so we strengthen the healthcare systems and availability of medicines.”

> Key Informant 19

> “You know half of Kenyans go to the private sector for their healthcare. So, we have to include private sector in UHC and all levels of private sector; the private hospitals, the private labs, the private pharmacies. So, we cannot force UHC models on people. It has to fit into the behavior of the clients of healthcare.”

> Key Informant 1

> “The private sector is a very big player in aspects like local production. You know, other than a long time ago when we had Dawa Pharmaceuticals which was government which was trying to produce essential medicines for the government, right now we don’t have any government led manufacturer of health products in the Country, they are all private sectors. I think private sector delivers around 40% of healthcare in the Country and all that is private. So, there is a very big role of the private sector especially in supporting the UHC agenda.”

> Key Informant 7

### Policy Recommendations

Our study outlines several policy recommendations where action can be taken to promote and ensure availability of essential medicines during novel times such as with the COVID-19 pandemic.

### Strengthening financial systems and policies of essential medicines

Sufficient investments in financing of essential medicines by the government is key in promoting its availability and ensuring the attainment of health goals such as the UHC. One of the ways can be through promoting innovative ways to enhance financial management of essential medicines funding by both local and international partners, hence ensuring sustainability. For instance, setting up an emergency kitty targeted at supporting acquisition, transportation and distribution of essential medicines is one of the ways. Additionally, to achieve UHC, a key factor is how the government finances essential medicines. The Country should aim to increase its budgetary allocation and ring fencing it to prioritize acquiring essential medicines during emergencies. As a long-term strategy, formulating and embedding policies that promote price regulation of essential medicines can promote availability and accessibility among the population. This can also reduce instances of catastrophic expenditure on medicines especially from impoverished households.

### Increasing resilience of supply chain and procurement systems of essential medicines

The COVID-19 pandemic underscored the significance of setting up both local and international procurement and supply chain systems that are both well-coordinated and adaptable to shocks that health systems can experience. Inefficiencies in supply chains can result in shortages and even wastages of essential medicines. Although other private and faith-based suppliers of essential medicines exist in Kenya, the bulk of the procurement is done by KEMSA, which serves majority of public institution and the population (majority of who are poor). The Country witnessed corruption and lack of transparency at KEMSA which marred the procurement, distribution and use of essential and non-essential medicines and products during the COVID-19 pandemic. To avert such future inefficiencies, the Country can adopt a mix of public, private and faith-based/NGO supply systems that can be complementary and ensure procurement and distribution of essential medicines even in times of crisis. Harmonization and collaboration by these sectors can reduce stock-outs and optimize availability of essential medicines through ways such as through pooled procurement.

### Strengthening legislation and regulatory policies around essential medicines

Pharmaceutical laws and regulations, create a basis under which operations such as manufacturing, trade and use of essential medicines are safeguarded. Whereas weak and conflicting policies can result in inefficiencies, responsive policies can help promote standards that will ensure availability of essential medicines. The pandemic exposed some of the weaknesses of regulatory policies in the Country such as; inadequate legal framework, conflicting policies and over-regulation of the pharmaceutical sector that hindered availability and accessibility of essential medicines. To avert future shortcomings, Kenya should invest in its regulatory authority institutions to promote proper industry regulation. This will ensure that stringent measures are undertaken in production, marketing, distribution, prescription and use of essential medicines in a bid to ensure progressiveness in the health of its population.

### Increasing domestic pharmaceutical manufacturing capacity

The pandemic presented a need for countries to foster their in-house production of essential medicines. With many high-income countries focusing on the needs and protection of their immediate population at the time, the fragility of global cooperations was exposed when faced by adversities. Many LMICs experienced further threats due to their heavy reliance on HIC for active pharmaceutical ingredients or finished products and the high burden of communicable and noncommunicable diseases. Therefore, to safeguard its population, Kenya must scale up and invest in its production and manufacturing portfolio of essential medicine and products to promote self-sufficiency. Crucial to this will be investing in the Country’s technology, infrastructure and human resource capacity which can promote local production. Additionally, incentivizing local production such as through establishment of conducive legislative framework increase local production. These legislations can include price controls, tax relief for manufacturers and overall establishment of good manufacturing practice (GMP) that attract local and international investments in production.

### Training of healthcare workforce for pharmaceutical production

Adequate investment in the right number and skill mix of pharmaceutical human resource can help transform the country’s pharmaceutical industry. Even though there has been an increase in pharmaceutical training institutions, the Country must prioritize expanding the training scope to not only focus on the clinical practice, but also to target other aspects such as manufacturing of complex molecules. Additionally, with the constant change in the pharmaceutical setting due to the emergence of new diseases (such as COVID-19) and changes in demographic landscape, a multi-dimensional approach is needed to align to these trends. This can be done through continuous professional training to ensure that the training content matches the industry requirements.

### Increase investments in the health information system for essential medicines

Kenya has made major strides on the health information system front in ensuring essential medicines’ availability. Albeit challenges, innovative ways such as online prescription and dispensing of drugs have been witnessed. These were seen to promote availability of drugs during the pandemic especially due to initial policies that saw restriction of movement, both of people and goods. The Country should expand its investments on health information system such as through digitization hence promote efficiency and effectiveness of services related to essential medicines availability.

### Conclusion

In conclusion, our study has identified several policy recommendations to promote and ensure the availability of essential medicines, particularly during novel situations like the COVID-19 pandemic. Firstly, strengthening the financial systems and policies related to essential medicines is crucial. This includes increasing investments in the financing of essential medicines by the government, both through innovative funding mechanisms and increased budgetary allocations. Formulating and implementing policies that promote price regulation of crucial medicines can also improve availability and accessibility, especially for vulnerable populations. Secondly, there is a need to enhance the resilience of the supply chain and procurement systems for essential medicines. This involves establishing well-coordinated and adaptable procurement and supply chain systems, both locally and internationally, that can effectively respond to shocks and emergencies. Collaboration between public, private, and faith-based/NGO supply systems can reduce stock-outs and ensure the availability of essential medicines even in crises. Thirdly, strengthening legislation and regulatory policies around essential medicines is essential. Adequate legal frameworks, harmonization of policies, and robust regulatory authority institutions are necessary to ensure the proper production, marketing, distribution, prescription, and use of essential medicines. Responsive policies can help promote standards that guarantee the availability of essential medicines.

Additionally, increasing domestic pharmaceutical manufacturing capacity is crucial for self-sufficiency in essential medicines. Investing in technology, infrastructure, and human resources and incentivizing local production through supportive legislative frameworks can promote local manufacturing and reduce dependency on external suppliers. Lastly, increasing investments in the health information system for essential medicines, such as digitization, can improve the efficiency and effectiveness of services related to availability and accessibility. Implementing these policy recommendations will promote the availability of essential medicines, ensure the population’s health and well-being, and build resilience in times of crisis.

## Data Availability

This was a qualitative study and all the data sets have been used in the analysis presented. The study does NOT Involve human subject tissue or trial at all.

